# Exploring the role of social networks in shaping STI disclosure among adolescents and young people using a health behavioral model in Lusaka, Zambia

**DOI:** 10.64898/2026.09.14.26362515

**Authors:** Gracious Witola, Sophie Inambwae, Adam Silumbwe, Steve Belemu, Gideon Phiri, Lucheka Sigande, Helen Ayles, Musonda Simwinga, Bernadette Hensen, Mwelwa M Phiri

**Affiliations:** Zambart House, School of Medicine, University of Zambia, Ridgeway, Lusaka, Zambia; Sexual and Reproductive Health Group, Department of Public Health, the Institute of Tropical Medicine, Antwerp, Belgium; Department of Clinical Research, London School of Hygiene and Tropical Medicine, London, United Kingdom; Department of Health Policy and Management, School of Public Health, University of Zambia, Lusaka, Zambia

**Author notes:** **Corresponding Author**: (GW).

## Abstract

Disclosure of sexually transmitted infections (STIs) is a key strategy for STI prevention and control. However, for adolescents and young people (AYP) aged 15-24, disclosure remains a challenge, usually shaped by several dynamics that affect AYP’s choice of whether to disclose or not, and to whom. This study sought to understand what influenced AYP decisions to disclose their STI status and to whom, using the Capability, Opportunity, and Motivation-Behavior (COM-B) model, in Lusaka, Zambia. This qualitative study, conducted from April-August 2024, included twenty in-depth interviews with AYP who participated in a cross-sectional STI prevalence survey, including those diagnosed with an STI, self-reporting STI symptoms, and reported ever being treated (regardless of symptoms). Additionally, five focus group discussions (FGDs) with AYP and another six FGDs with community members were conducted. Data were then analysed thematically using the COM-B model. The capability to disclose was influenced by AYP’s limited knowledge about STIs. Physical opportunity to disclose was shaped by the availability and accessibility of youth-friendly Sexual and Reproductive Health (SRH) services, while community and self-stigma, and relationships and household dynamics also shaped AYP’s social opportunities to disclose their STI status. Motivation to disclose was influenced by concern for one’s overall health, individual values and beliefs, and the need to seek support when experiencing an STI. However, fear of negative reactions reduced AYP’s motivation and willingness to disclose their STI status. AYP’s decisions to disclose an STI diagnosis is shaped by a network of personal and social influences. Social support plays an important role in disclosure, with supportive social networks facilitating the process and thus encouraging individuals to seek treatment. STI interventions should not only focus on individuals but also on increasing STI awareness, reducing stigma, and leveraging existing social support within the broader community context.

## Introduction

The four leading curable sexually transmitted infections (STIs), chlamydia trachomatis, Neisseria gonorrhoeae, trichomonas vaginalis, and syphilis, collectively pose a significant public health burden. According to the World Health Organization (WHO), each day more than one million people aged 15–49 acquire curable STIs worldwide [1], with sub-Saharan Africa bearing the highest burden of STIs [2,3]. STI control remains a major public health priority, as untreated STIs can lead to severe health consequences, including infertility, stillbirths, increased HIV transmission risk, psychological distress, and even death [3–6].

Adolescents and young people (AYP) aged 15-24 are particularly vulnerable to STIs, with AYP residing in Southern African countries experiencing a disproportionately high prevalence of STIs compared to older adults [7–10]. This elevated vulnerability is linked to factors such as early sexual debut, sex with multiple partners, condomless [11–13], and transactional sex [14] that are often driven and/or compounded by substance abuse, physical and sexual abuse [12]. Other factors include stigma around young people’s sexuality [14], and limited access to sexual and reproductive health (SRH) services [15].

Much progress has been made in the detection and provision of effective treatment for common STIs including through the availability of low-cost rapid diagnostics, single-dose regimens of antibiotics and antivirals, particularly in high burden, low resource settings[16]. Despite these improvements, AYP who have a confirmed STI diagnosis or STI-like symptoms often do not disclose, which negatively impacts prevention, management and uptake of STI treatment [17]. AYP often choose not to disclose a confirmed STI diagnosis or STI-like symptoms due to a complex interplay of factors, including fear of rejection or violence, pervasive stigma, feelings of embarrassment, a perceived lack of obligation to disclose, [5,7,18,19], and deeply ingrained sociocultural beliefs that often confine sexual health discussions to adulthood or within the context of marriage [7,20]. In addition, AYP may avoid healthcare facilities and hesitate to disclose their STI-like symptoms to healthcare providers due to concerns about confidentiality, judgmental attitudes, and limited STI literacy [7].

In 2022, a scoping review of local evidence on health and HIV among AYP by UNICEF-Zambia, highlighted how navigating STI disclosure among AYP remains a major challenge, largely because we do not clearly understand AYP’s decision making regarding whether to disclose or not, to whom, and in what context[21]. Most local studies on STI disclosure tend to focus on HIV [22–25] and specifically partner notification [5,19]. Additionally, the literature on STI disclosure pays limited attention to AYP disclosure within broader social support networks such as health providers, parents, friends, or relatives [5], and offers little insight into the social factors shaping this disclosure process [21]. This is particularly important because the disclosure of STIs for AYP, unlike for adults, requires that AYP navigate several health systems and social dynamics [26–28]. Moreover, depending on whom AYP disclose to and get advice from, this may influence their decisions regarding treatment and health practice.

This study sought to understand what influenced AYP decisions to disclose a confirmed STI diagnosis or STI-like symptoms, and to whom, using the COM-B model, in Lusaka, Zambia. The capability, opportunity, and motivation-behaviour (COM-B) model [29] provides a useful lens for understanding the multi-level factors shaping AYP’s decision-making process on whether to disclose or not, a confirmed STI diagnosis or STI-like symptoms within their social networks (Fig 1).

**Fig 1.**
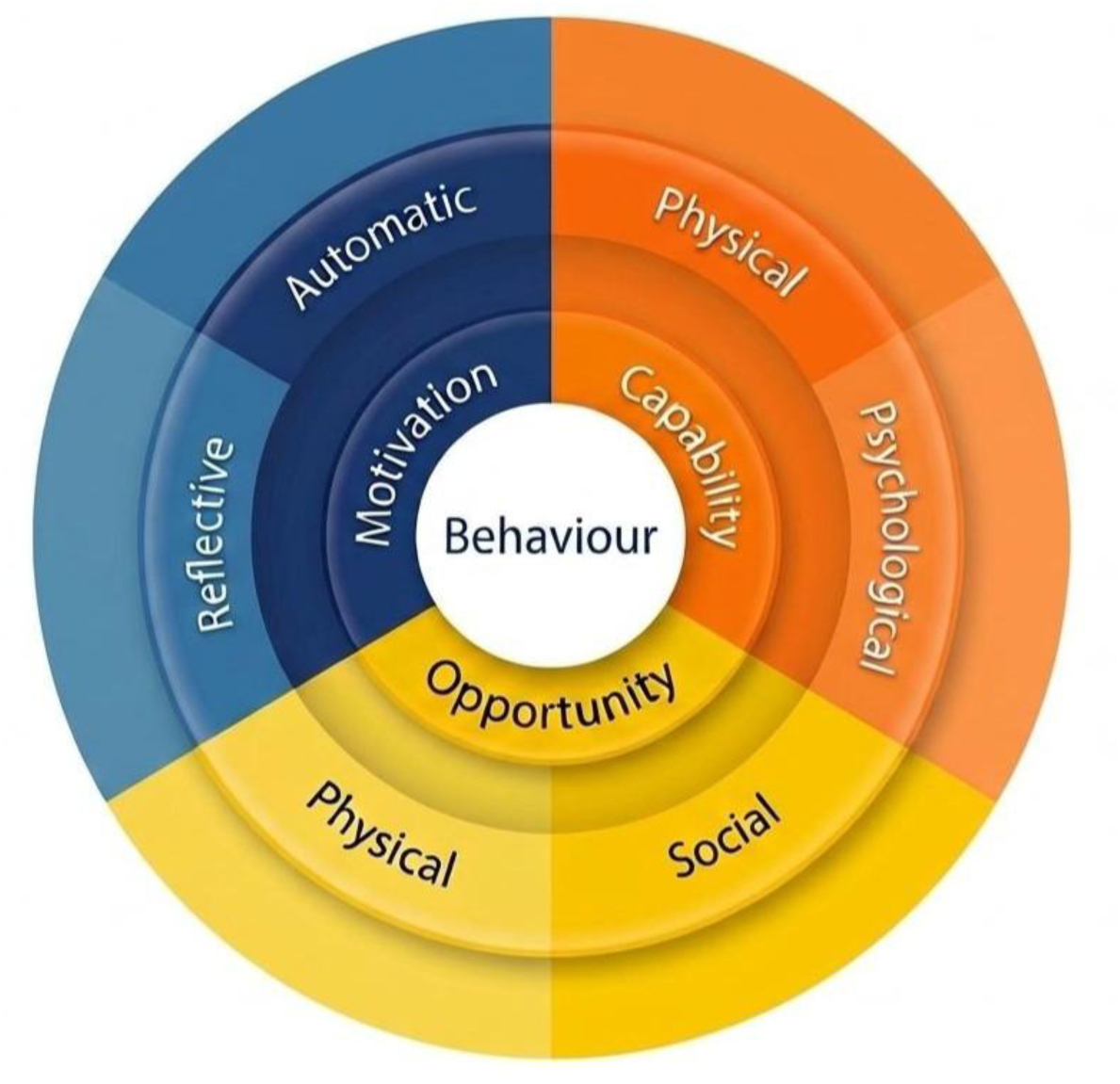
Capability, Opportunity, Motivation-Behavior (COM-B) model of behavior. Reproduced from Michie et al. [30].

### Study framework

The COM-B model provides a structure to comprehend why and how certain behaviours concerning AYP STI disclosure occur, and therefore a basis for those who wish to intervene to change the behaviour. It argues that, for behaviours to occur, three important factors must be present–capability, opportunity and motivation–which interact overtime resulting in a particular behaviour that becomes part of a dynamic system with positive and negative feedback loops [30]. The greater the capability and opportunity, the more likely one is motivated to engage in a particular behaviour. Capability comprises the psychological (Knowledge) and physical (skills) attributes that make behaviour possible. Opportunity is the attribute of the external environment that allows or prompts an individual to behave in a certain way, including physical opportunities (location and resources) and social opportunities (social support and cultural norms). Motivation includes reflective and automatic internal processes that energise and direct behaviour, such as beliefs and emotions. The COM-B model is the core part of the Behaviour Change Wheel (BCW), which articulates interventions and policy options that can be adopted to change behaviour. The BCW uses the COM-B model to understand behaviour and then helps choose the right strategies to change it [29].

## Methods

### Study design

This was an exploratory qualitative study that was nested within a bigger cross-sectional survey, “Understanding the prevalence of and care seeking pathways for sexually transmitted infections among adolescents and young people aged 15-24 in Lusaka, Zambia”, that sought to estimate the prevalence of curable STIs among adolescent girls and young women (AGYW), and adolescent boys and young men (ABYM) aged 15 to 24 year in Lusaka, Zambia. In this paper, we report on data from the two phases of this qualitative research: a formative phase to inform the design of the main survey and a phase when qualitative data were collected in parallel to the survey.

### Study setting

We conducted this study in a high-density, urban community situated in the south-west of Lusaka, Zambia. The study community has a population of approximately 200,000 people based on the 2022 census [31]. It is also serviced by a Level 1 referral hospital and four health posts with various ethnic groups, predominantly speaking Nyanja.

### Population, sampling and recruitment

The study population included the AYP, community members, community leaders, and STI service providers. All participants were selected based on their age, gender, being a guardian of an AYP, provision of STI services, and leadership in the community. The in-depth interview (IDI) participants–AYP–were randomly selected from the STI database of the main study by (LS) using Stata and then located in their households by the research assistants (RA). Focus group discussion (FGD) participants were purposively sampled with the help of the main project’s community engagement (CE) team (SB and GP), the community advisory board members and the local health facility. These individuals were familiar with the community through active outreach in public spaces such as markets, schools, churches, and recreational facilities (such as football pitches and drinking places). Participants were purposively sampled to ensure a variation among AYP, guardians of AYP, and STI service providers across geographic zones within the community.

### Data collection

We collected data using IDIs and FGDs conducted between April and August 2024. Both the IDIs and FGDs guides were developed by the research team based on the literature review. The FGD interview questions focused on understanding of STIs, perceptions about STIs among AYP, notification of partners and parents/guardians, and community support, while the IDIs included additional questions on personal STI symptoms, testing experiences, and access to care. The data were collected by the first author (GW), a female social scientist with an MA in social science and experience in designing and conducting qualitative research, with the support of two trained male researchers and facilitators (SB and GP). All data were collected in Bemba, English or Nyanja, depending on participant preference, to facilitate ease of communication. The total number of IDIs and FGDs that were conducted was guided by the principle of theoretical saturation, we stopped conducting additional interviews when we noticed no new information from additional data [32].

#### Focus group discussions

During the formative phase (April and May 2024), we conducted five (n=5) FGDs with adolescent girls, adolescent girls and young women (AGYW), adolescent boys, adolescent boys and young men (ABYM), and a combined group of AYP (Table 1). Alongside the survey conducted between June and August 2024, six (n=6) focus group discussions (FGDs) were conducted with community members, community leaders, young adults, and STI service providers. Among the community members, two FGDs were conducted with caregivers of AYP, one with men and another with women who had AYP under their care. Another two FGDs were conducted with young adults and STI service providers, respectively (Table 1). Lastly, two FGDs were conducted with community leaders, including traditional healers, community zonal leaders, health– and development-related committee leaders, teachers, and church leaders. All FGDs were held in the health facility youth-friendly spaces where participants felt safe to be in. Refreshments and a meal were given as compensation (Table 1).

**Table 1.** Categories of participants, demographics and data collection methods.

| Methods | Participant Type | Gender |  | Total |
| --- | --- | --- | --- | --- |
|  |  | M | F |  |
| IDIs | AYP self-reporting STI-like symptoms either with accessing or not accessing care | 4 | 4 | 8 |
|  | AYP who reported ever being treated regardless of symptoms | 2 | 2 | 4 |
|  | AYP diagnosed with an STI during survey | 4 | 4 | 8 |
| FGDs | All adolescents and young people (AYP) aged 15-24 | 4 | 5 | 9 |
|  | Adolescent girls (AG) aged 15-17 |  | 9 | 9 |
|  | Adolescent girls and young women (AGYW) aged 18-24 |  | 12 | 12 |
|  | Adolescent boys (AB) aged 15-17 | 8 |  | 8 |
|  | Adolescent boys and young men (ABYM) aged 18-24 | 11 |  | 11 |
|  | Community members (men) | 9 |  | 9 |
|  | Community members (women) |  | 8 | 8 |
|  | Community leaders 1 | 4 | 6 | 10 |
|  | Community leaders 2 | 5 | 4 | 9 |
|  | Young adults aged 25 to 35 | 4 | 5 | 9 |
|  | Service providers | 2 | 8 | 10 |

#### In-depth interviews

To understand pathways to care and factors affecting AYP’s access to health services when experiencing STI-like symptoms, we conducted 20 IDIs with AYP aged 15–19 and 20–24 years who self-reported STI-like symptoms, reported ever being treated, or were diagnosed with an STI through testing performed as part of the survey (see Table 1). The IDIs took place either at the facility’s youth-friendly space or in the participants’ homes. Refreshments and a meal were given as compensation.

### Data management and analysis

All IDIs and FGDs were audio recorded. Field notes were also taken using a template and then typed in Microsoft Word immediately after data collection. Audio recordings were transcribed verbatim and translated from local languages to English during the transcription process. GW reviewed the transcriptions for accuracy and quality against the audio recordings. We used both deductive and inductive thematic analysis [33]. The main themes were deductively derived from the study framework, including the factors shaping AYP behaviours–capability, motivation and opportunity. Emergent themes were inductively derived from context-specific issues shaping AYP disclosure in social networks that arose from the field notes and detailed reading of the transcripts. The analysis process started with GW reading through field note reports and a sample of six transcripts to identify and define codes, then develop a draft code-list, with the initial main and sub-themes, which was later discussed and modified with the two Principal Investigators (PIs) who are joint last two authors (BH) and (MMP) and the research team. GW then proceeded to manually code the rest of the transcripts, often meeting with the research team to discuss and update the code-list (Table 2). After the coding was done, GW developed code reports for all themes.

**Table 2.** Qualitative data code-list.

| **COM-B Dimension** | **Main themes** | **Sub-themes** |
| --- | --- | --- |
| Psychological capability | ▪ Limited knowledge about STIs among AYP | ▪ Misinformation, myths, and misconceptions of STI |
| Social and physical opportunity | ▪ Social Stigma | ▪ Community and self-stigma |
|  | ▪ Relationship and household dynamics | ▪ Availability of trust between the AYP and the person they disclose to<br>▪ Quality, length, and nature of relationship with sexual partner<br>▪ Absence of a supportive disclosure environment in the household |
|  | ▪ Availability of adolescent SRH services | ▪ Youth friendly spaces enable AYP's disclosure but not widely used |
| Reflective and automatic motivation | ▪ Concern for one's overall health | ▪ Desire to prevent STIs and seek prompt treatment for those infected AYP |
|  | ▪ Individual values and beliefs | ▪ Moral obligation to disclose to sexual partner |
|  | ▪ Soliciting support | ▪ A need for emotional and financial support for treatment and recovery from an STI |
|  | ▪ Fear | ▪ Negative reactions by partners, family, friends and community |

### Ethical Considerations

We obtained Ethical approvals for the study from the University of Zambia Biomedical Research Ethics Committee (REF.No.4805-2024), the London School of Hygiene and Tropical Medicine Ethics Committee (REF: 29990), the Institute of Tropical Medicine institutional review board (REF.1748/24), the Ethics committee of the University Hospital of Antwerp (ID 6436), and the Zambian National Health Research Authority (REF:NHREB005/25/03/2024). During the survey, participants were informed about the possibility of being contacted for follow-up IDIs, and they consented to this. Those who were later randomly selected from the STI database, provided consent, and were formally contacted to schedule the IDIs. Written informed consent was obtained from AYP aged 18–24 years, while written parental consent along with assent was obtained for 15–17-year-old adolescents. Participants also verbally consented to having their interviews and FGDs audio recorded. To maintain confidentiality, we anonymised the participants using participant identification numbers.

## Results

The findings below are organized thematically, based on the three components of the COM-B model: capability, opportunity and motivation with one to four sub-themes each.

### Psychological capability

In this domain, findings showed that psychological capability to disclose a confirmed STI diagnosis or STI-like symptoms was influenced by limited knowledge driven by misinformation, myths, and misconceptions.

#### Misinformation, myths, and misconceptions

Aside from knowing the names of the two common STIs, syphilis and gonorrhea, being aware of common STI symptoms, such as genital sores, the consequences of STIs, such as infertility, and treatment such as ‘Benzathine Benzylpenicillin’; most AYP had limited knowledge of STIs or were misinformed. For example, a young person explained how he only got to know about one STI during the survey.

> *“One thing I have observed is that we are short of information … it’s not there in the community. Even myself I have only heard today about trico vaga (trichomonas vaginalis) what is it?” [Man_ 25-29 years].*

Most AYP were unsure of the symptoms attached to specific STIs, and some did not always understand what they were suffering from and were unaware they had an STI, making it difficult for them to disclose to anyone. For instance, a male AYP diagnosed with chlamydia narrated how he held off disclosing his diagnosis to his girlfriend.

> “… *not yet I haven’t told her because the results came whereby, I didn’t know where I got it (Chlamydia) from till yesterday when I was researching everything” [Young man_ 20-24 years].*

AYP seemed to find it easier to disclose if they convinced themselves that the STI-like symptoms they were experiencing were unrelated to the common STIs mentioned earlier. In fact, for many AYP any STI-like symptoms were assumed to be a fungal infection linked to shared toilets or poor hygiene, which facilitated disclosure to immediate relations without hesitation. An adolescent diagnosed with an STI had this to say.

> *“I thought it was a fungal infection (AYP diagnosed with syphilis). I told my mom. My mother is also not cognizant with these issues of STIs, so she just thought it was fungal, so she went and bought medicine for fungal.” [Adolescent girl_ 15-19 years].*

In addition, study participants mentioned that community misconceptions around treatment and prevention of confirmed STI or STI-like symptoms also encouraged non-disclosure. AYP believed that washing their private parts with alcohol after sex, warm lemon water, aloe vera, garlic, or roots would prevent or cure STI-like symptoms. These myths and misconceptions discouraged disclosure as AYP would hold it out not until the symptoms became severe, and they had to go to the facility for treatment. An adolescent during an FGD had this to say.

> *“Three quarters use roots when they have STIs; very few go to the clinic for treatment. Some people put lemon in water and wash their private parts” [Adolescent boy_ 15-19 years].*

### Social and physical opportunity

This domain highlights participant views on factors that influence both social and physical opportunity for AYP to disclose a confirmed STI diagnosis or STI-like symptoms. Social opportunity to disclose was shaped by community and self-stigma, relationship and household dynamics, including the availability of trust between the AYP and the person they disclose to, the quality, length, and nature of the relationship with a sexual partner, and the absence of a supportive disclosure environment in the household. The availability and accessibility of youth-friendly SRH services also influenced AYP’s physical opportunity to disclose a confirmed STI diagnosis or STI-like symptoms.

#### Community and self-stigma

Stigma was cited by most participants as a major barrier to the disclosure of STIs among AYP. Most AYP narrated that being diagnosed with an STI would make them feel embarrassed and disappointed in themselves, culminating in their failure to accept their situation. Such stigma stemmed from community misconceptions that individuals diagnosed with an STI were ‘dirty’, which made AYP feel ashamed, discouraging disclosure.

> *“Syphilis ‘ni doti’ (it’s dirty) if one has sex with a man and does not bathe, they can get syphilis” [Young woman_ 20-24 years].*

Community members and AYP perceived that stigma was also driven by the symptoms that accompany certain STIs such as bad odor or sores on one’s private parts. Such symptoms would result in AYP having to avoid others or vice versa. The church was also seen as influential in the AYP disclosure process within the community. Sexual topics were viewed as offensive or taboo, with sex before marriage, considered sinful or associated to demons. This caused some community members to shun AYP with STIs, further reinforcing non-disclosure.

> *“… they (church) don’t touch on subjects related to HIV and STIs, to them, those are … those are demons … People are dying in churches, we see the coffins, but they are still living in denial saying it cannot happen to someone filled with the Holy Spirit” [Man_ 60-64 years].*

#### Availability of trust between the AYP and the person they disclose to

According to the participants, AYP would only disclose that they were experiencing symptoms or had a confirmed STI to someone they trusted or had a relationship with. The participants felt that AYP usually disclosed to their partners or any other person so that they could be advised on how to deal with STI symptoms. The trusted individuals included partners, elderly persons within the family, community health workers and even close friends that would guide the AYP on the different courses of action including buying off-the counter or traditional medication.

> *“We also get information from the elderly who know about STIs. They can tell when one has an STI because of experience, these elderly with grey hair provide suggestions for traditional medicine ‘yachiboyi’” [Young woman_ 20-24years]*
>
> *“So, they could choose who to tell based on how much they trust them with regards to confidentiality” [Adolescent girl_15-19 years]*

#### The quality, length, and nature of relationship with a sexual partner

Participants narrated that AYP who were in committed loving relationships were more likely to disclose to their partners compared to those with difficult and unloving partners. The length of the relationship with a partner also played a key role in AYP disclosure of STIs. Some participants explained that AYP in long-term relationships that have gone on to get married were more likely to disclose if they had an STI.

> *“It depends on the person you are with. Others are supportive and can even go with you so that you both get tested. While others are not” [Young Woman_ 20-24 years]*

In contrast, those AYP starting new relationships were less likely to disclose due to fear of being viewed as promiscuous or losing the relationship altogether.

> *“It depends on how comfortable one is with their partner. If you are not comfortable, you cannot tell them. I did not tell my partner because it was our first-time having sex, then I tell him I am experiencing these symptoms” [Adolescent girl_ 15-19 years].*

The nature of the relationship also played a role in disclosure. Some participants stated that AYP with multiple sexual partners, were less likely to disclose that they had an STI due to not knowing who they acquired the STI from or feared being labelled as promiscuous by their partners and others. Regarding casual sexual partners, AYP were said to be less obliged to disclose because they were not their primary partner. One young person had this to say about disclosing to multiple partners.

> *“But how, where do you start from if you have about 7 partners. It is better you don’t tell them or tell the one you love” [Young woman_20-24].*

In contrast, some AYP argued that they would confront their secondary partners, disclose their positive STI test results, and request that they also do a test to establish who infected the other if they believed they were the ones who had been infected.

> *“I was from having sex with them and there was no one else, it was her. So, she is the one that I followed and told her. She started saying her ‘apologies.’ I went to shout at her, saying, you are stupid. I did not tell my main girlfriend because I knew that she was not the one that had given this to me” [Adolescent boy_ 15-19].*

#### Absence of a supportive disclosure environment in the household

Most participants explained that many parents did not provide a supportive environment for AYP to disclose their confirmed STI diagnosis or STI-like symptoms in the household, with parents considered strict and difficult to talk to. This was mainly because it was culturally inappropriate for parents to talk to their children about sexual issues. Participants relayed the need for parents to be more open and build relationships with their children as AYP could disclose and could learn about STIs from their parents. A service provider had this to say during an FGD.

> *“Aah the knowledge gap that she talked about is probably because of culture because rarely do parents talk about STIs to their children. They feel it could be a taboo.” [Man_ 45-49 years].*

#### Youth friendly spaces enable AYP’s disclosure but not widely used

Some participants felt the health facility, particularly the youth friendly space, supported AYP disclosure by providing information on STIs, including on symptoms and treatment. However, many noted that some AYP were reluctant to use these spaces due to social judgment, confidentiality breaches, lack of understanding of how the health system worked or what to expect during a visit to one.

> *“Uh, it is not every adolescent in the community that knows about the youth-friendly space and most people that come here are those that are in school. If you did it in the community, you would be able to capture even those that are out of school and those that are in marriages” [Woman_ 25-29 years]*.
>
> *“They want to but are usually scared because they don’t know what the procedures will look like” [Adolescent Boy_ 15-19years].*

In addition, costs of certain tests equipment, and medication in government facilities further discouraged disclosure, leading many AYP to prefer visiting pharmacies rather than queue at the health facility.

> *“They go to the drugstore because the hospital usually prescribes medication, so rather than them queuing up at the hospital they go straight to the drugstore” [Young woman_ 20-24 years].*

### Reflective and Automatic Motivation

Motivation was influenced by concern for one’s overall health, including the desire to prevent STIs and seek prompt treatment for AYP infected with an STI, a moral obligation to disclose to sexual partners, the need for emotional and financial support for treatment and recovery from an STI, and fear of negative reactions from partners, friends, and close relations.

#### Desire to prevent STIs and seek prompt treatment for those infected AYP

Many AYP viewed the concern for their overall health status as a key factor shaping the STI disclosure process. Some AYP considered it important to disclose to protect their partner (s) from acquiring the infection and the possible death that could result from untreated STIs like syphilis. Others felt the need to disclose STI-like symptoms or confirmed STI diagnosis to sexual partners (s) for them to test for STIs, or access treatment and prevent reinfection.

> *“It is important because if you realize when it is too late you can die. You can also protect your partner from contracting the STI” [Young Man_ 20-24 years].*

However, several participants also noted a lack of concern for overall health among some AYP. This was often expressed through phrases like “I don’t care what happens.” Some AYP even ignored STI symptoms or felt the health problem would disappear on its own. Others only decided to go to the health facility when the symptoms became serious, delaying testing and disclosure.

> *“A few tests, because a lot have many girls and say even if I am with it, it is fine to let me die…There is no need for me to test” [Young man_ 20-24 years].*

#### A moral obligation to disclose to sexual partners

Individual values and beliefs among AYP were noted to play a key role in disclosure of STIs and care seeking. Some AYPs believed that they had a moral obligation to disclose their STI status or symptoms, particularly to their sexual partners. However, the participants mentioned that it was easier for AYP to disclose a negative result than a positive one. Indeed, some waited to see their test results or after treatment before disclosing to anyone.

> *“I told him when I was cured.” [Adolescent girl_ 15-19 years]*.
>
> *“There is that girl you like people, I can tell her out of the rest” [Adolescent boy_ 15-19 years].*

#### A need for emotional and financial support for treatment and recovery from an STI

Some AYPs were motivated to disclose a confirmed STI diagnosis or STI-like symptoms to solicit emotional and financial support from their partners, parents and friends. Emotional support included being encouraged to get tested individually or with one’s partner, being escorted to the clinic or advised on where to get treatment and adhering to medication. Financial support on the other hand included buying of medication and other supplies. Similarly, some AYPs disclosed to close friends, neighbors, or relatives to get support or advice on where to access treatment.

> *“…The advantage that I found in informing my mother, was that she bought me the medication I needed … she’d ask me if I have taken my medication when she knocks off.” [Young woman_ 20-24 years].*

#### Fear of negative reactions by partners, friends and close relations

Most participants stated that the fear of the negative reactions of partners, parents, and close relations to disclosure was a barrier. Some AYP had concerns about being perceived as promiscuous, rejection, gossip, and embarrassment which could potentially affect one’s reputation and the chances of future relationships. Some AYP, especially AGYW, feared violence from their sexual partners due to gender imbalances and being blamed for infecting their partners.

> *“People don’t say they fear being left, being embarrassed, or being gossiped about to others or to future potential partners” [Young woman_ 20-24 years]*
>
> *They are scared that they will be beaten; many don’t disclose. The men are even more free to disclose after they have been treated and have been cured. That’s when they disclose to their women about the infection and confront them” [Man_ 25-29 years].*

Some participants mentioned fear of the pain from the STI injection, especially ‘Benzathine Benzylpenicillin’ for syphilis, and fear of a positive result as barriers to disclosure as some AYP would refuse to disclose to health personnel or get tested due to this. One participant shared this during an IDI.

> *“I did not want to go to the clinic … well … I thought they’d tell me I had more diseases in me, so that’s why others say it’s better for them not to know how their health is” [Young man_ 20-24years].*

## Discussion

This study sought to understand what influenced AYP decisions to disclose their STI status and to whom, using the COM-B model, in Lusaka, Zambia. The capacity of AYP to disclose was influenced by limited knowledge about STIs while the opportunity to disclose was shaped by availability and accessibility of youth-friendly SRH services. Socially, community and self-stigma, relationships, and household dynamics also shaped AYP opportunities to disclose their STI status. Concern for one’s overall health, individual values and beliefs and the need to solicit support when someone has an STI motivated AYP disclosure. However, fear of negative reactions influenced the desire for AYP to disclose that they had an STI.

Our findings show that limited information in the form of myths and misconceptions about STIs influenced AYP’s decision to disclose a confirmed STI diagnosis or STI-like symptoms. Although AYP were aware of gonorrhea and syphilis, they had limited knowledge of other STIs with most unsure of what they were suffering from or assuming their symptoms were due to a fungal infection. Similar findings were reported in a study in Ghana [34] among early adolescents, where participants were mainly aware of HIV and gonorrhea but had limited knowledge of other STIs. Unlike our study, adolescents in Ghana were aware of most STIs and did not confuse them with a fungal infection. This could possibly be because the study was conducted in schools, where they are more likely to receive comprehensive sexuality education compared to AYP in our community-based sample. However, our findings align with a study in Uganda [35] which found that although adolescents recognized sexual activity as a mode of STI transmission, many also believed STIs were transmitted through sharing toilets or poor hygiene. These misconceptions may cause AYP to downplay STI-like symptoms and delay disclosure. This misconception also extends to treatment practices, highlighting the need for interventions that strengthen comprehensive sexuality education not only among AYP, but also within households, schools, and communities.

Beyond the transmission of misconceptions regarding STIs, our findings show the dual role of social networks in shaping disclosure among AYP. AYP were more likely to disclose in environments they perceived as safe and supportive. Similarly, a systematic review on STI disclosure to sexual partners[19] found that disclosure was influenced by a partner’s supportiveness. A study in the United States [5] reported that AYP with supportive and open parents were more likely to seek and receive emotional, informational, and financial support, including accompaniment to health facilities and treatment support, while strict or conservative parents discouraged disclosure. Similarly, a study conducted in Zambia among young people living with HIV [22] found that supportive and caring home environments increased confidence in disclosing HIV status to others.

Fear of gossip, stigma, and negative reactions emerged as major barriers to STI disclosure among AYP. Similar findings from studies in Zimbabwe [7,36] among young people aged 16-24 reported that fear of being perceived as promiscuous, blamed for the infection, or socially rejected due to damaging rumors spreading within the community, including among family members, discouraged disclosure to partners, parents, and peers. Fear of partner violence and harsh parental reactions, particularly among AGYW, further increased reluctance to disclose [7,36] highlighting the influence of power dynamics within relationships. In Uganda [35] the visibility of STI symptoms, such as sores or odors, among adult men and women (≥18 years) intensified stigma. Similarly, a study in South Africa [37] among teenagers aged 13–19 found that religious and cultural norms that framed sexual health discussions as immoral, limited education and support for AYP, especially unmarried young people. These findings highlight the need for comprehensive sexuality education and stigma reduction interventions that extend beyond schools and health facilities into families, communities, and places of worship. Community-based interventions that address harmful gender norms and misconceptions about STIs may also help reduce fear and discrimination associated with disclosure.

Social dynamics such as the length and nature of relationship influenced STI disclosure among AYP as found in previous studies on disclosure to sexual partners [18,19]. Disclosure was less likely in casual or non-committed relationships, where participants often felt it was unnecessary, and more likely among primary or long-term partners where closeness and trust had been established [19]. This may reflect limited trust and fears of damaging rumors that could lead to social rejection and harm to one’s reputation. Future community interventions should address approaches supporting disclosure across these contexts. In contrast, some participants prioritized a sense of moral obligation to disclose despite these fears, consistent with findings from Australia among people aged 15–29 in Victoria [18].

Health system challenges were also reported to influence STI disclosure among AYP. Most participants felt that communities lacked youth-friendly SRH services that promote STI awareness, testing, and treatment, ultimately affecting STI diagnosis and disclosure. Similarly, a scoping review in Zambia [21] reported that despite government efforts to promote disclosure through youth-friendly services, major challenges remain, including limited accessibility, concerns about healthcare provider attitudes and confidentiality, and service hours that conflict with school schedules. Some AYP described how limited understanding of available services reduced their confidence to seek care and disclose symptoms. Studies among AYP in Zambia [38] and adolescents in Indonesia [39] suggest that these barriers extend beyond STI care to other sexual and reproductive health services. Limited access to youth-friendly and trusted health services may reduce AYP’s willingness and ability to disclose STIs. Community-based STI awareness programmes and improving young people’s understanding of available services may also increase confidence in navigating the healthcare system and seeking timely care.

### Strengths and limitations

This study had several limitations. First, although it explored STI disclosure, participants were not specifically recruited based on prior disclosure, which may have limited insights into factors that support disclosure. Second, the COM-B model was applied during analysis rather than informing the design of data collection tools, potentially limiting the depth of behavioral exploration. Further, while the model may have shown clear distinctions among the themes, this was not the case, highlighting the complexity and interconnectedness of factors that influence AYP decisions to disclose an STIs. Despite these limitations, the study had key strengths. Participants were randomly selected from AYP who had experienced STI-like symptoms or had been diagnosed with an STI, allowing for the inclusion of lived experiences. The study also included both girls and boys, as well as parents and caregivers, providing broader perspectives on STI disclosure.

## Conclusion

Our findings show that the decision to disclose an STI is shaped by a network of personal and social influences. Social support can play an important role in disclosure of STIs, with supportive social networks facilitating disclosure process thus encouraging AYP to seek timely treatment. STI interventions should focus not only on individuals but also on increasing STI awareness and reducing stigma within the broader community context. Further research is needed to explore interventions that effectively improve STI disclosure and healthcare navigation among AYP in low-resource settings.

## Data Availability

All data produced in the present study are available upon reasonable request to the authors.

## Acknowledgements

Sincere thanks to the study community leaders, the study participants, and other stakeholders who supported this work.

